# Data-efficient prostate cancer classification from T2-weighted MRI with mechanistic auditing and a dual-agent quality-control workflow

**DOI:** 10.64898/2026.09.20.26363529

**Authors:** Vahid Monfared, Mohammad Hadi Gharib, Reza Rawassizadeh

## Abstract

Diffusion-weighted images are sometimes unavailable or degraded in prostate MRI. We asked whether T2-weighted MRI alone retains usable cancer-related signal and whether the computation can be audited. An image-level transfer-learning ensemble reached out-of-fold AUC 0.883 (95% CI 0.836–0.924) on 243 de-identified institutional T2 images; a handcrafted image-level model reached 0.924 (0.889–0.953) and, in an exploratory external test of 137 Prostate158 patients, 0.643 (0.545–0.734) against 0.494 previously. A volume-level DINOv2-attention model reached AUC 0.717 (0.572–0.853) in 133 held-out biopsy-linked patients, 0.733 with test-time augmentation, and a locked precursor 0.618 (0.566–0.668) in 561 independent PROMIS patients. Distillation reproduced the frozen teacher with *R*² 0.776, a transparent boosting machine 0.678 and a readable model tree 0.550, exposing dependence on gland size, low T2 signal and texture. A bounded dual-agent application gates input quality and anatomy before frozen inference, returning audit metadata. These findings support further evaluation of auditable T2-only analysis when complementary sequences are unavailable or degraded.

## Introduction

Multiparametric MRI has reshaped the diagnostic pathway for prostate cancer. In PROMIS, MRI before biopsy detected clinically significant cancer far more often than systematic transrectal biopsy^1^, and in PRECISION an MRI-targeted pathway found more significant disease while diagnosing less insignificant disease^2^. PI-RADS version 2.1 defines interpretation as a multi-sequence task in which T2-weighted imaging provides the anatomy and the dominant transition-zone score, while diffusion-weighted imaging dominates the peripheral zone^3^. We accept that standard; our question is different. How much cancer-related morphological information does the T2 series carry by itself, can it be learned from a few hundred examples, does it survive a move to other institutions, and can the model be made transparent?

This matters practically, because the complementary sequences are not always usable whereas T2 acquisition is standard and reproducible across centers. Among 11,319 prostate MRI examinations, 4.8% of patients had a hip arthroplasty, and moderate to severe susceptibility artefact on diffusion images was associated with a 26% lower cancer detection rate^4^; when rectal gas degraded diffusion images, detection rates were preserved, which the authors attributed partly to compensation by T2-weighted and contrast-enhanced images^5^. Such failure modes motivate T2-only models as sequence-resilient research and triage tools, and support faster pathways where scanner time, contrast agents and subspecialty reading are scarce. They are not intended for autonomous screening.

Two obstacles stand in the way. Current systems are data-hungry: bi-parametric networks needed well over 2,000 training examinations to approach expert performance^6^, and the PI-CAI system that outperformed 62 radiologists was developed within 10,207 examinations^7^. Most hospitals cannot assemble that scale. Models are also opaque: a heat map shows where a network responds but not how a probability is assembled, and post-hoc explanation is weaker evidence than examining the computation itself^8^. Knowledge distillation^9^ offers a route from a black box to a glass box that can be read and tested.

We address both obstacles within one auditable framework (Fig. 1). Self-supervised representations^10^ and T2-specific transfer learning reduce the parameters estimated locally, while nested out-of-fold evaluation limits optimistic internal estimation. A volume-level attention model^11^ is locked before evaluation in a non-overlapping UCLA cohort, and a separately locked precursor is evaluated once in the independent PROMIS cohort. Distillation converts the frozen teacher into a high-fidelity neural student, transparent additive models, explicit equations and readable rules, and a bounded two-agent application checks input compatibility and quality before returning the frozen prediction with its audit state. Our earlier preprint classified individual T2 slices and showed near-chance transfer^12^; the present work advances to patient-level evaluation and computational auditing.

**Fig. 1.**
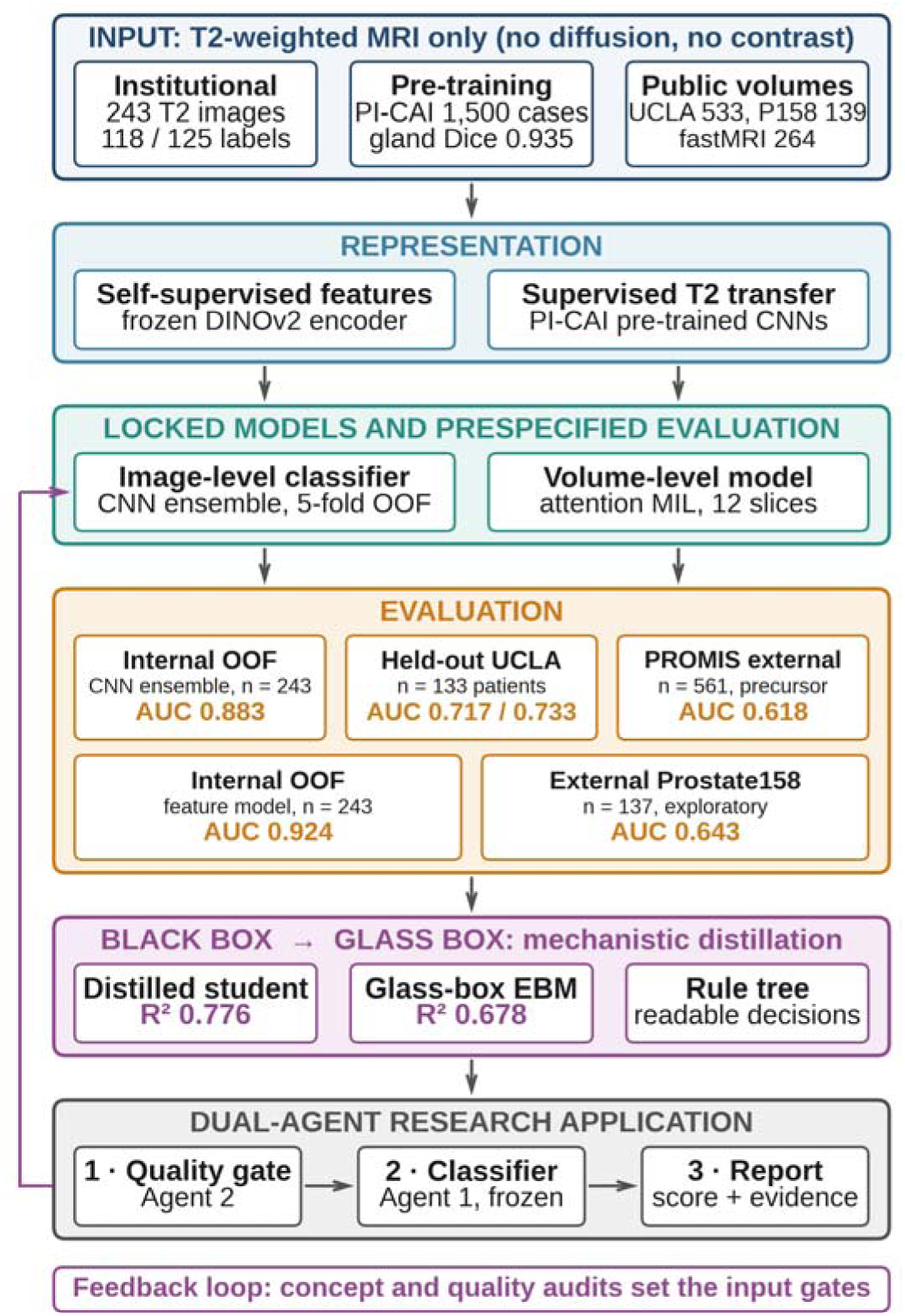
The framework, from T2-only input to an audited report. The only image input is the T2-weighted series, and most capacity comes from frozen self-supervised and T2-specific pre-training. Evaluation is reported at three levels that are never pooled: internal out-of-fold estimation, a prespecified non-overlapping held-out analysis in UCLA patients, and one independent evaluation of a locked precursor in PROMIS patients. The lower evaluation row is a separate image-level feature model trained on the institutional images alone; its Prostate158 value is exploratory. Distillation converts the frozen network into a student model, ablation-tested concepts and readable rules. In the application the quality agent (Agent 2) runs first and can warn, reject or abstain; only then does the classifier agent (Agent 1) run the frozen model, and the purple loop shows the audit feeding back into the input gates. MIL, multiple-instance learning; OOF, out-of-fold.

## Results

### Study cohorts

Table 1 lists every data source by unit of analysis. The institutional archive held 268 exported axial T2-weighted images; hashing removed 17 duplicates and 8 images with colored annotation overlays were excluded, leaving 243 de-identified images, 118 cancer and 125 normal. Patient identifiers and examination-level linkage were not retained, so unique patient and examination counts are unavailable. Public data served four roles: T2 pre-training and gland segmentation (1,500 PI-CAI examinations), volume-model development (539 patients: 400 UCLA, 139 Prostate158), the interpretability audit (264 fastMRI Prostate patients), and two evaluation cohorts never used for fitting, 133 non-overlapping UCLA patients (113 cancer, 20 benign) and 561 PROMIS patients^1^.

**Table 1.** Data sources, units of analysis and roles.

| Cohort | Role in this study | Unit and size | Reference standard |
| --- | --- | --- | --- |
| Institutional, Golestan University of Medical Sciences | Development and out-of-fold validation of the image-level classifier | 243 de-identified images (118 cancer, 125 normal); patient and examination counts unavailable in the analytic export | Institutional cancer or normal label supplied with the dataset |
| PI-CAI (public) | T2 pre-training and gland segmentation | 1,500 examinations; 4,500 axial T2 slices | Case labels; whole-gland masks |
| Prostate158 (public) | Volume model development; exploratory external test of the separate image-level model | 139 patients (83 with tumor, 56 without) | Expert tumor annotation |
| UCLA Prostate-MRI-US-Biopsy (public) | Volume model development | 400 patients (304 cancer, 96 benign) | Targeted and systematic biopsy |
| UCLA Prostate-MRI-US-Biopsy (public) | Non-overlapping held-out test, locked before model fitting | 133 patients, 133 MRI studies (113 cancer, 20 benign) | Biopsy, any cancer (primary) |
| PROMIS cohort (public trial data)<sup>1</sup> | Independent external stress test of a locked precursor model | 561 patients, one prediction each | Biopsy; any cancer and Gleason score $\geq 7$ |
| fastMRI Prostate (public) | Mechanistic distillation | 264 patients, development split | Teacher probability only |
| Prior reader study | Human T2-only reading | 22 cases, 5 radiologists | Pathology-confirmed cancer (8) or normal (14) |

Note: Cohorts are reported separately because they differ in unit of analysis, reference standard and prevalence. Neither evaluation cohort was used for training, tuning, calibration or threshold selection. The held-out cohort shares a public source with development data; the PROMIS cohort does not.

### Internal performance from T2-weighted images only

Across 243 out-of-fold predictions, each from a fold model that never saw the image, the ensemble reached ROC-AUC 0.883 (95% CI 0.836-0.924) and average precision 0.840 (Fig. 2a,b and Table 2). At the locked threshold of 0.441 it classified 101 of 118 cancer and 101 of 125 normal images correctly: sensitivity 0.856, specificity 0.808, positive predictive value (PPV) 0.808, negative predictive value (NPV) 0.856, accuracy 0.831, F1 0.831 and Brier 0.148 (Fig. 2c,d). Fold AUCs ranged from 0.883 to 0.997 (Supplementary Fig. 1 and Supplementary Table 1), an independently developed hybrid reached 0.881 on the same images (Supplementary Note 1), and net benefit exceeded biopsy-all and biopsy-none from 0.05 to about 0.80 (Supplementary Fig. 2).

**Fig. 2.**
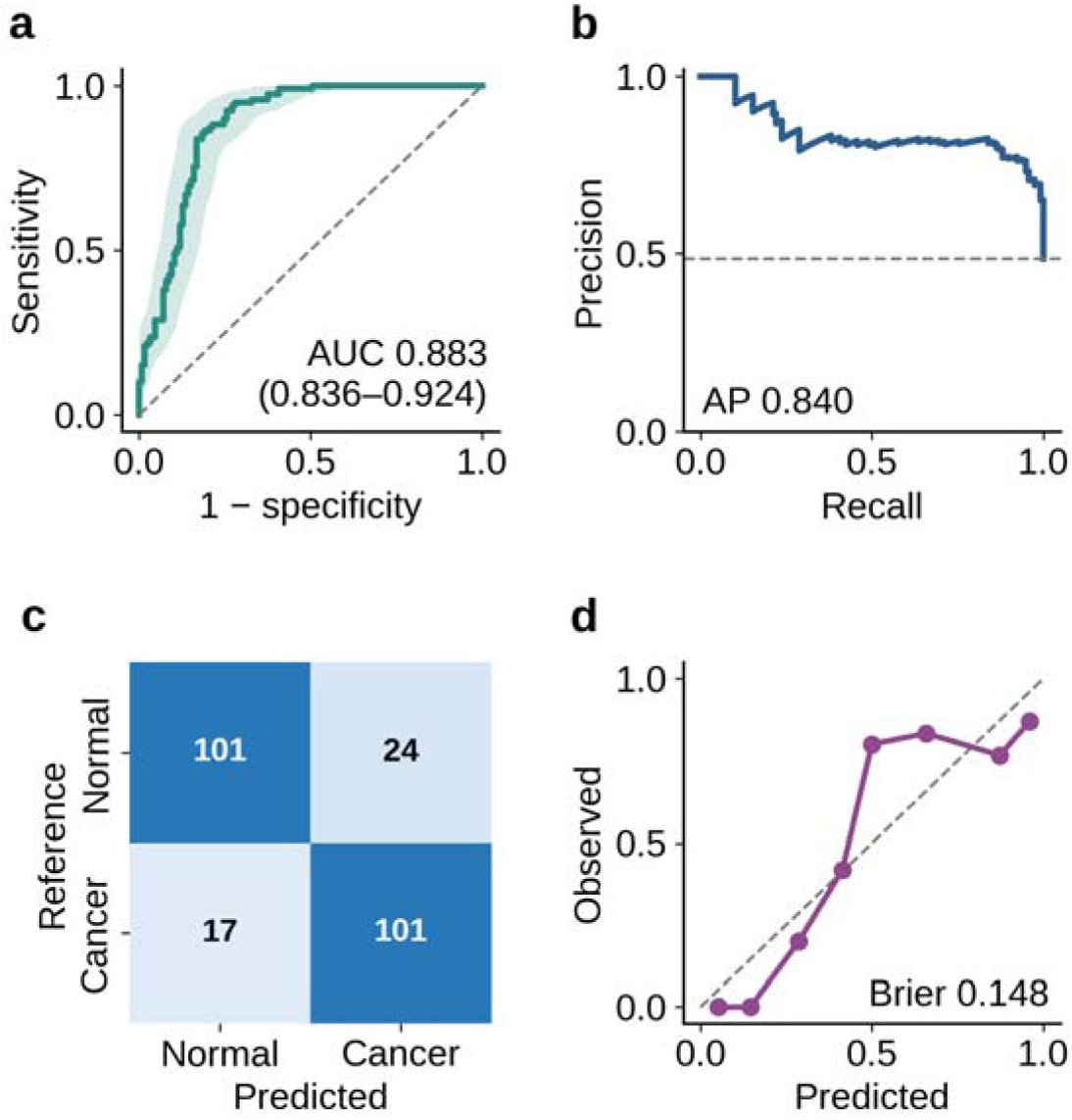
Internal performance on 243 institutional T2 images. Every value comes from pooled five-fold out-of-fold predictions, so no image contributed to the model that scored it. **(a)**, Receiver operating characteristic curve with the pointwise 95% bootstrap band (2,000 resamples); the model separates cancer from normal images with an AUC of 0.883. **(b)**, Precision–recall curve; the dashed line is the cancer prevalence of 0.486, and precision stays near 0.80 across most of the recall range. **(c)**, Confusion matrix at the locked threshold of 0.441, showing balanced errors (24 false positives, 17 false negatives). **(d)**, Calibration in eight quantile bins: predicted probabilities track observed risk at the extremes and understate risk near 0.5, which is why the threshold rather than the raw probability defines the operating point. AP, average precision.

**Table 2.**
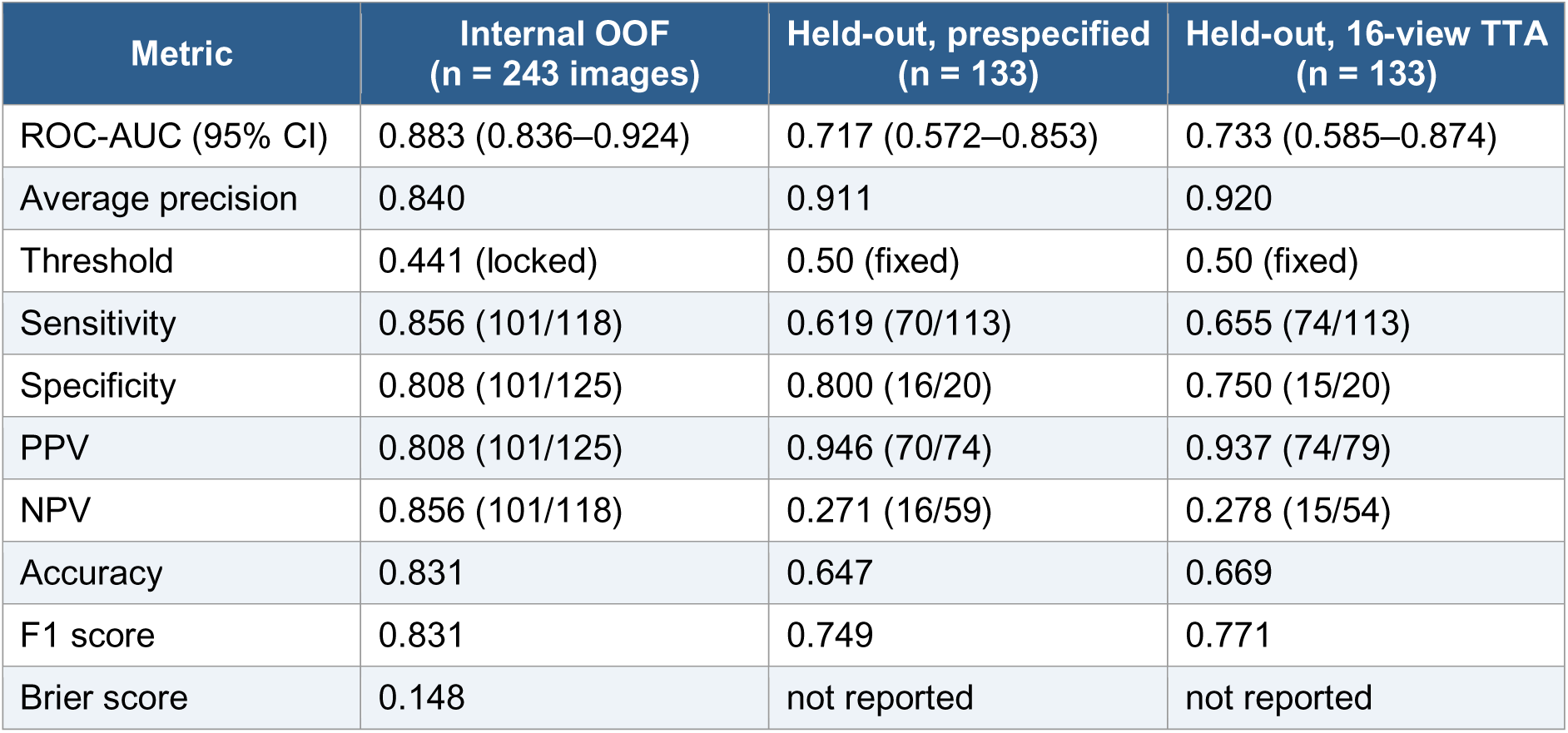
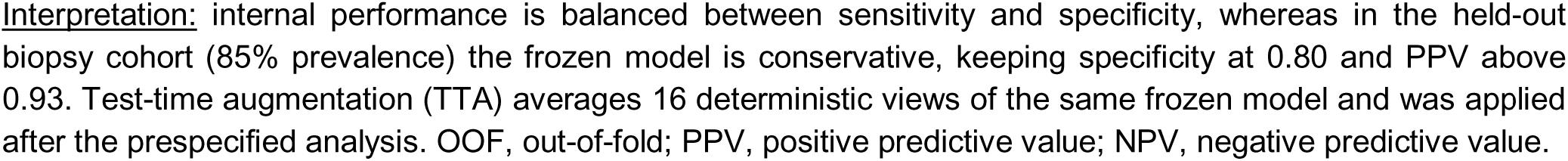
Discrimination and operating-point performance of the locked models.

| Metric | Internal OOF<br>(n = 243 images) | Held-out, prespecified<br>(n = 133) | Held-out, 16-view TTA<br>(n = 133) |
| --- | --- | --- | --- |
| ROC-AUC (95% CI) | 0.883 (0.836–0.924) | 0.717 (0.572–0.853) | 0.733 (0.585–0.874) |
| Average precision | 0.840 | 0.911 | 0.920 |
| Threshold | 0.441 (locked) | 0.50 (fixed) | 0.50 (fixed) |
| Sensitivity | 0.856 (101/118) | 0.619 (70/113) | 0.655 (74/113) |
| Specificity | 0.808 (101/125) | 0.800 (16/20) | 0.750 (15/20) |
| PPV | 0.808 (101/125) | 0.946 (70/74) | 0.937 (74/79) |
| NPV | 0.856 (101/118) | 0.271 (16/59) | 0.278 (15/54) |
| Accuracy | 0.831 | 0.647 | 0.669 |
| F1 score | 0.831 | 0.749 | 0.771 |
| Brier score | 0.148 | not reported | not reported |

Interpretation: internal performance is balanced between sensitivity and specificity, whereas in the held-out biopsy cohort (85% prevalence) the frozen model is conservative, keeping specificity at 0.80 and PPV above 0.93. Test-time augmentation (TTA) averages 16 deterministic views of the same frozen model and was applied after the prespecified analysis. OOF, out-of-fold; PPV, positive predictive value; NPV, negative predictive value.

### Held-out biopsy-linked testing

Institutional images were exported slices, whereas public cohorts provide complete volumes, so a volume-level model was built for patient-level testing: a frozen DINOv2 encoder^10^ with gated attention multiple-instance learning^11^, developed on UCLA and Prostate158 patients^13,14^. A non-overlapping subset of 133 UCLA patients was fixed by manifest before fitting and excluded from training, calibration and threshold selection. For the prespecified any-cancer endpoint, ROC-AUC was 0.717 (95% CI 0.572–0.853), rising to 0.733 (0.585–0.874) with 16-view test-time augmentation, with average precision 0.911 and 0.920 (Fig. 3a and Table 2). At the fixed threshold of 0.50 the model identified 70 of 113 cancers and 16 of 20 benign biopsies: sensitivity 0.619, specificity 0.800, PPV 0.946, NPV 0.271, accuracy 0.647 and F1 0.749 (Fig. 3b,c and Supplementary Figs. 3 and 4); the high PPV and low NPV follow from the 85% prevalence of this biopsy population. Because development and testing drew on the same public source, this is a held-out test, not independent external validation, and with only 20 benign biopsies the interval is wide.

**Fig. 3.**
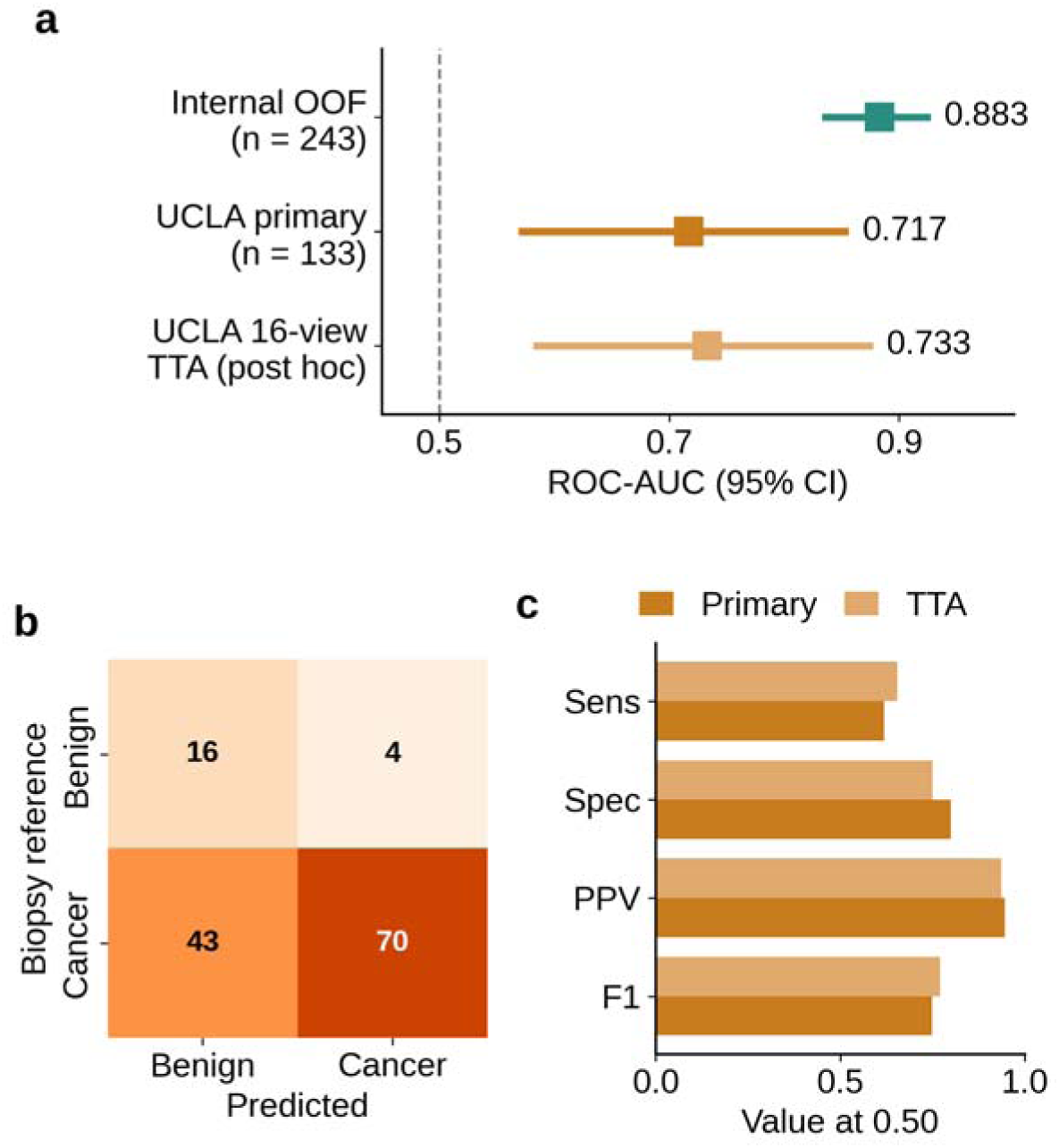
Held-out biopsy-linked evaluation of the locked T2 volume model in 133 non-overlapping patients. **(a)**, ROC-AUC with 95% bootstrap confidence intervals for the internal out-of-fold estimate and for the two held-out analyses of the frozen model; all three intervals lie above chance (dashed line). **(b)**, Confusion matrix of the prespecified analysis at the fixed threshold of 0.50 against the biopsy reference: 70 of 113 cancers and 16 of 20 benign biopsies were correctly assigned. **(c)**, Operating-point metrics for the prespecified analysis and for 16-view test-time augmentation (TTA). Cancer prevalence was 85.0%, which raises positive predictive value (PPV 0.946 and 0.937) and lowers negative predictive value (0.271 and 0.278), so a positive score is far more informative than a negative one here. Sens, sensitivity; Spec, specificity. Because development and testing drew on the same public source, this is a held-out test; the independent external result is reported separately in Table 3.

**Fig. 4.**
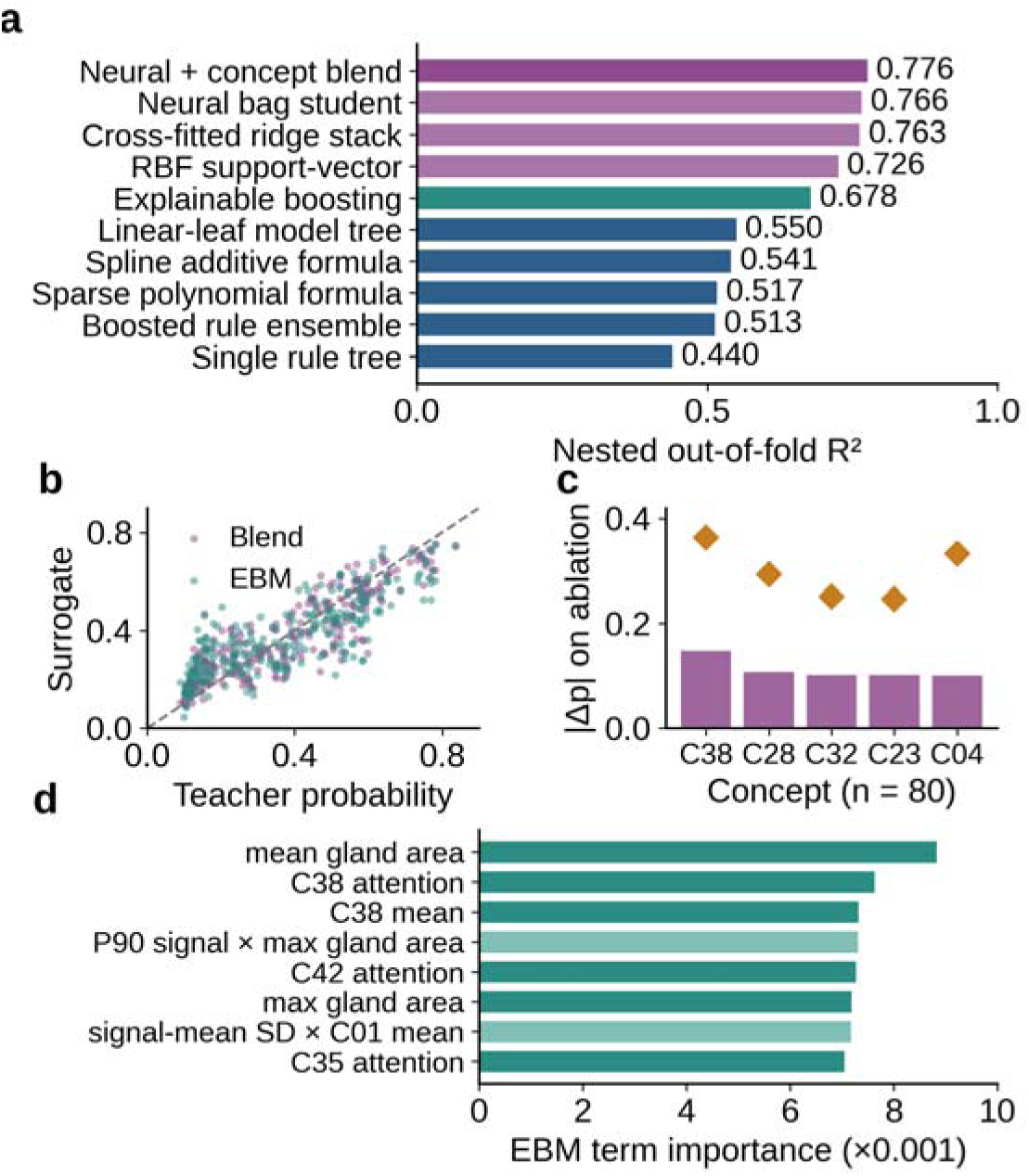
Computational interpretability of the locked volume model (264 fastMRI development patients). **(a)**, Fidelity of eleven surrogate configurations under five-fold nested cross-validation: purple, high-capacity students; green, the intrinsically transparent explainable boosting machine (EBM); blue, compressed glass-box descriptions. Fidelity falls in an ordered way as the description becomes more readable. **(b)**, Out-of-fold surrogate predictions against teacher probabilities for the 70:30 blend and the EBM; the dashed line is identity. **(c)**, Mean (bars) and maximum (diamonds) absolute probability change after zero-ablating the five most influential sparse-autoencoder concepts in 80 cases. **(d)**, The eight strongest EBM terms: solid bars are single-feature shapes, pale bars pairwise interactions.

**Table 3.** Patient-level evaluation and comparison with the previous external benchmark.

| Evaluation | Cohort and unit | Model version | Size | ROC-AUC (95% CI) |
| --- | --- | --- | --- | --- |
| Previous framework<sup>12</sup> | Prostate158, slice level | Best of six deep models | 19 cases, 142 slices | 0.494 (range 0.416–0.494) |
| This study, held-out test | UCLA, patient level, any cancer | Final model, frozen before testing | 133 patients | 0.717 (0.572–0.853) |
| This study, held-out test with TTA | UCLA, patient level, any cancer | Final model, 16 views | 133 patients | 0.733 (0.585–0.874) |
| This study, independent external | PROMIS, patient level | Locked precursor, applied once | 561 patients | Any cancer 0.618 (0.566–0.668); Gleason $\geq 7$ 0.607 (0.560–0.653) |
| This study, exploratory external | Prostate158, patient level, any cancer | Image-level model, institutional training only | 137 patients | 0.643 (0.545–0.734) |

**Interpretation:** the earlier slice-level evaluation transferred near chance, whereas the present patient-level analyses show moderate discrimination that is retained, in attenuated form, in a fully independent cohort. UCLA represents a patient-disjoint held-out analysis from the same public source; PROMIS is the only fully independent external cohort. Results should not be pooled because different locked model versions were evaluated, and no statistical ranking across cohorts is implied. In PROMIS, average precision was 0.792 for any cancer and 0.625 for Gleason score 7 or higher, with sensitivity 0.627, specificity 0.566 and accuracy 0.610. The Prostate158 row evaluates a separate image-level model trained on the institutional images alone, for which that collection is genuinely external; it is exploratory because several configurations were compared on it before the reported one was selected. TTA, test-time augmentation.

### Independent external testing under domain shift

A second evaluation used a cohort with no connection to model development. A separately locked precursor version of the volume-model pipeline was applied once to 561 patients of the PROMIS trial^1^, with no training, fine-tuning, recalibration or threshold adjustment and with predictions fixed before outcomes were read. For any biopsy-detected cancer, ROC-AUC was 0.618 (95% CI 0.566–0.668), average precision 0.792, sensitivity 0.627, specificity 0.566 and accuracy 0.610; for Gleason score 7 or higher, ROC-AUC was 0.607 (0.560-0.653) with average precision 0.625 (Table 3). Discrimination stayed above chance under a different scanner fleet, protocol, reference standard and case mix. Because a precursor was evaluated, this is an independent framework-level stress test, not external validation of the final model.

### Patient-level evaluation in context

Table 3 places the previous Prostate158 external evaluation alongside the present patient-level evaluations, preserving their methodological differences. In the earlier framework, six neural configurations produced slice-level AUCs of 0.416 to 0.494 on Prostate158^12^. Here the final volume model achieved AUC 0.717 in the prespecified UCLA held-out cohort and 0.733 with exploratory test-time augmentation, and a separately locked precursor achieved 0.618 in the fully independent PROMIS cohort. Because these analyses differ in cohort, model version, endpoint and unit of analysis, they provide complementary evidence of transportability rather than a statistical ranking. Bi-parametric systems using diffusion imaging have reported higher discrimination^7^ but address a different input setting from the T2-only fallback evaluated here.

Because the volume model was developed partly on Prostate158 patients, that collection cannot test it. A separate image-level model, trained on the 243 institutional images alone, was applied once to 137 Prostate158 patients. Patient-level ROC-AUC was 0.643 (95% CI 0.545-0.734), average precision 0.728, sensitivity 0.951 and F1 0.754 at the internally locked threshold (Table 3), the first estimate in this line of work whose interval excludes chance, against 0.416 to 0.494 previously^12^. It is exploratory: several representations and aggregation rules were compared on this cohort before the reported configuration was chosen (Supplementary Note 3).

### From black box to glass box: computational interpretability

The locked volume model acted as the teacher on 264 fastMRI development patients^15^; no outcome labels were used, and the teacher was never retrained. Eleven surrogate configurations were compared under five-fold nested cross-validation, all selection confined to training folds (Fig. 4a and Supplementary Table 2). The highest fidelity came from a fixed blend of the neural bag and concept students (Equation 1),

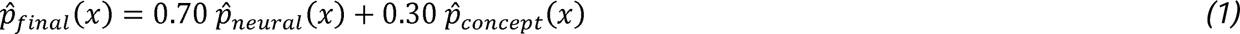

which reproduced teacher probabilities with *R*² 0.776, mean absolute error 0.074, Pearson r 0.881 and 86.0% agreement at the 0.50 cut-off (Fig. 4b). Among intrinsically transparent models the strongest was an explainable boosting machine (*R*² 0.678, mean absolute error 0.090, agreement 83.7%), which expresses every prediction as an exact sum of one-dimensional feature shapes and twenty pairwise interactions (Equation 2),

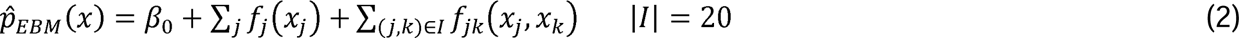

so, a case can be explained by listing the signed terms that produced its score. The largest contributions came from mean and maximum gland area, concepts C38, C42 and C35, and two interactions between signal statistics and gland size (Fig. 4d). More compressed descriptions lost fidelity in an ordered way: a model tree reached *R*² 0.550, a spline additive formula 0.541, a sparse polynomial 0.517, a boosted rule ensemble 0.513 and a decision tree 0.440. No surrogate reached *R*² 0.80, so we report the fidelity–readability frontier rather than its best point.

Removing the highest-attention slice changed the probability by a mean of 0.040, against 0.012 for the lowest (paired Wilcoxon *R =* 3.8 × 10⁻⁹; Supplementary Fig. 5). Zeroing concept C38 shifted it by a mean of 0.148 (maximum 0.363) in the same direction in all 80 audited cases (Fig. 4c and Supplementary Table 3). C38 tracked 10th-percentile signal intensity (r = −0.37), and C28 texture homogeneity (r = 0.50) and low grey-level co-occurrence contrast (r = −0.60), consistent with the radiological description of cancer on T2 images as low-signal tissue^3^. Blurring raised the teacher probability by a mean of 0.167 across 40 cases (Supplementary Fig. 6), a texture dependence the quality agent is designed to catch. These are computational dependencies of a frozen model, not biological mechanisms.

Two descriptions are directly readable (Fig. 5). The model tree splits once on the mean segmented gland area ā of a patient’s selected slices, then applies a transparent linear equation L in each branch (Equation 3),

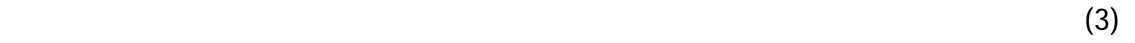

**Fig. 5.**
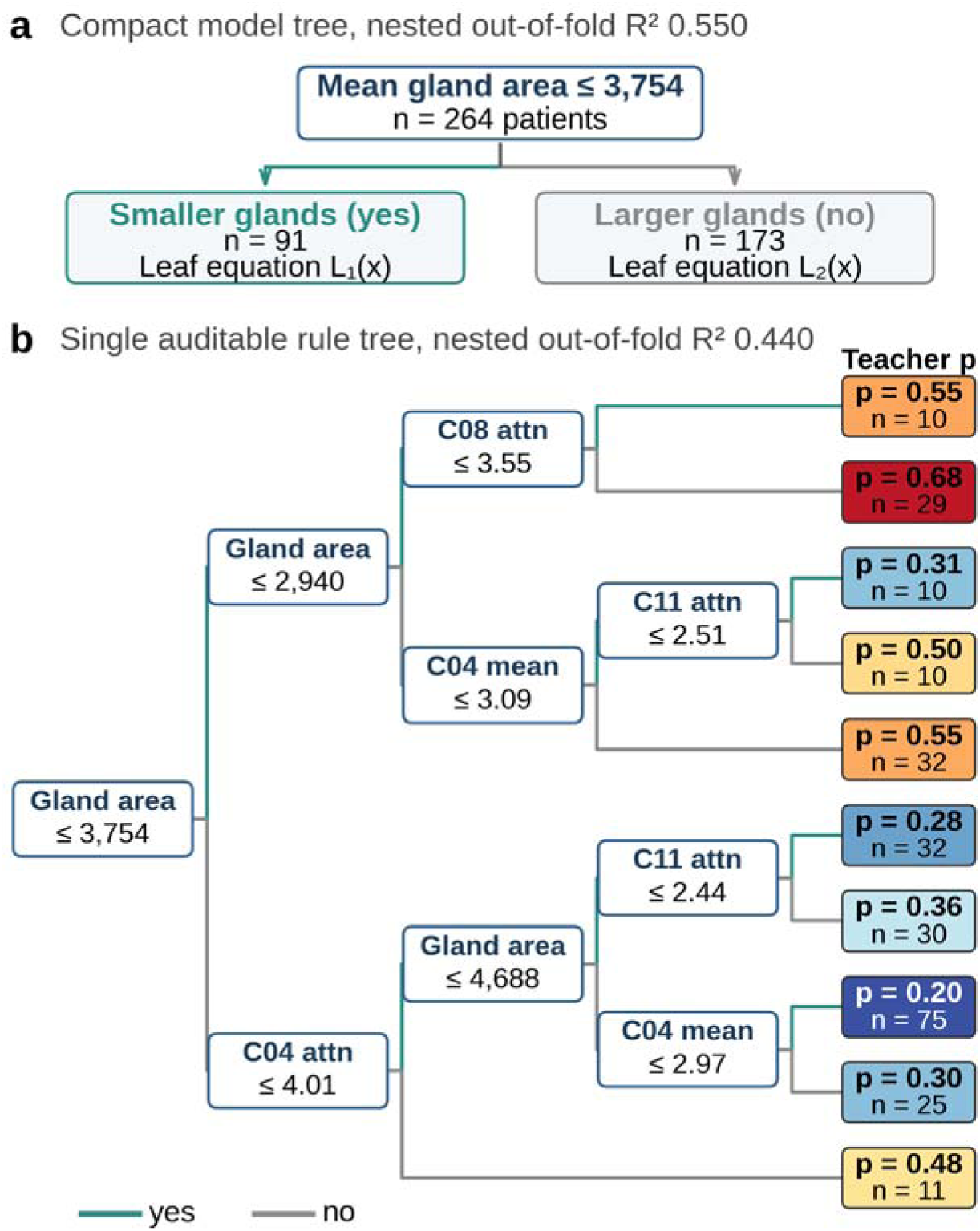
Readable rule structures distilled from the locked volume model. Both structures were fitted to teacher probabilities on 264 fastMRI development patients; teal marks the branch where a condition holds, grey where it does not. **(a)**, Compact model tree: one split on mean segmented gland area divides the cohort into 91 and 173 patients, and each branch applies its own linear equation (Equation 3; coefficients in Supplementary Fig. 7); nested out-of-fold fidelity 0.550. **(b)**, Single auditable rule tree ( 0.440), read left to right; terminal boxes give the mean teacher probability and the number of patients reaching that leaf, coloured from low (blue) to high (red). Cxx is a sparse-autoencoder concept, attn its attention-weighted activation and mean its average activation.

with 91 patients in the smaller-gland branch and 173 in the larger-gland branch; in both leaves the largest coefficient is the spread of the selected slice positions, followed by concept activations and gland-area terms (Supplementary Fig. 7 and Supplementary Note 2). The rule tree ( ) expands the same logic into ten rules: teacher probability was highest (0.68, 29 patients) in small glands with high C08 attention and lowest (0.20, 75 patients) in large glands with low C04 activity. Both views carry one message a radiologist can check: gland size sets the baseline, and concepts tracking low T2 signal and texture move the score up or down. Figure 6 draws the whole audit as one map.

**Fig. 6.**
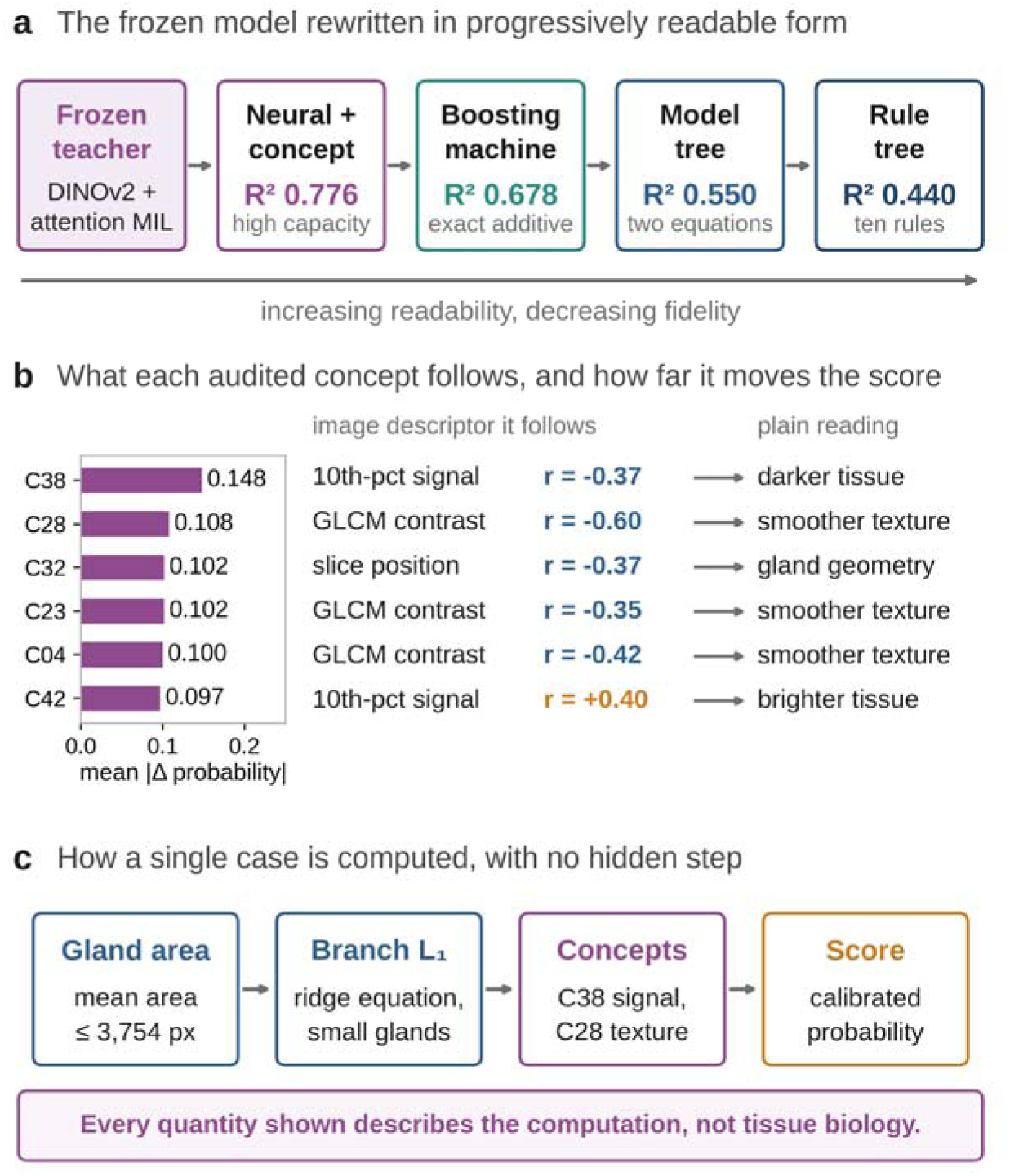
Mechanistic map of the audited computation. **(a)**, The frozen teacher rewritten in progressively readable form, with nested out-of-fold fidelity at each step; readability rises from left to right as fidelity falls, so the choice of surrogate is an explicit trade-off. **(b)**, The six most influential sparse-autoencoder concepts, the mean absolute probability change produced by zero-ablating each one in 80 cases, the image descriptor each concept follows and how that descriptor reads in the image. **(c)**, One case computed end to end: the gland-area split selects a branch, the branch equation and concept activations adjust the score, and the calibrated probability follows. All quantities describe the computation of a frozen model and are not claims about tissue biology.

### Dual-agent application

The framework is delivered as a public research application with two bounded roles (Fig. 7). The quality agent normalizes the upload, checks compatibility with an axial grayscale prostate T2 image, and may warn, reject or abstain. The classifier agent then runs the frozen ensemble and returns the calibrated probability with the locked threshold, fold agreement, an auxiliary prostate region estimate and a class-targeted Grad-CAM map^16^. The quality agent records notes but cannot alter the probability, calibration or threshold; every report carries the model version, threshold, image fingerprint and quality state.

**Fig. 7.**
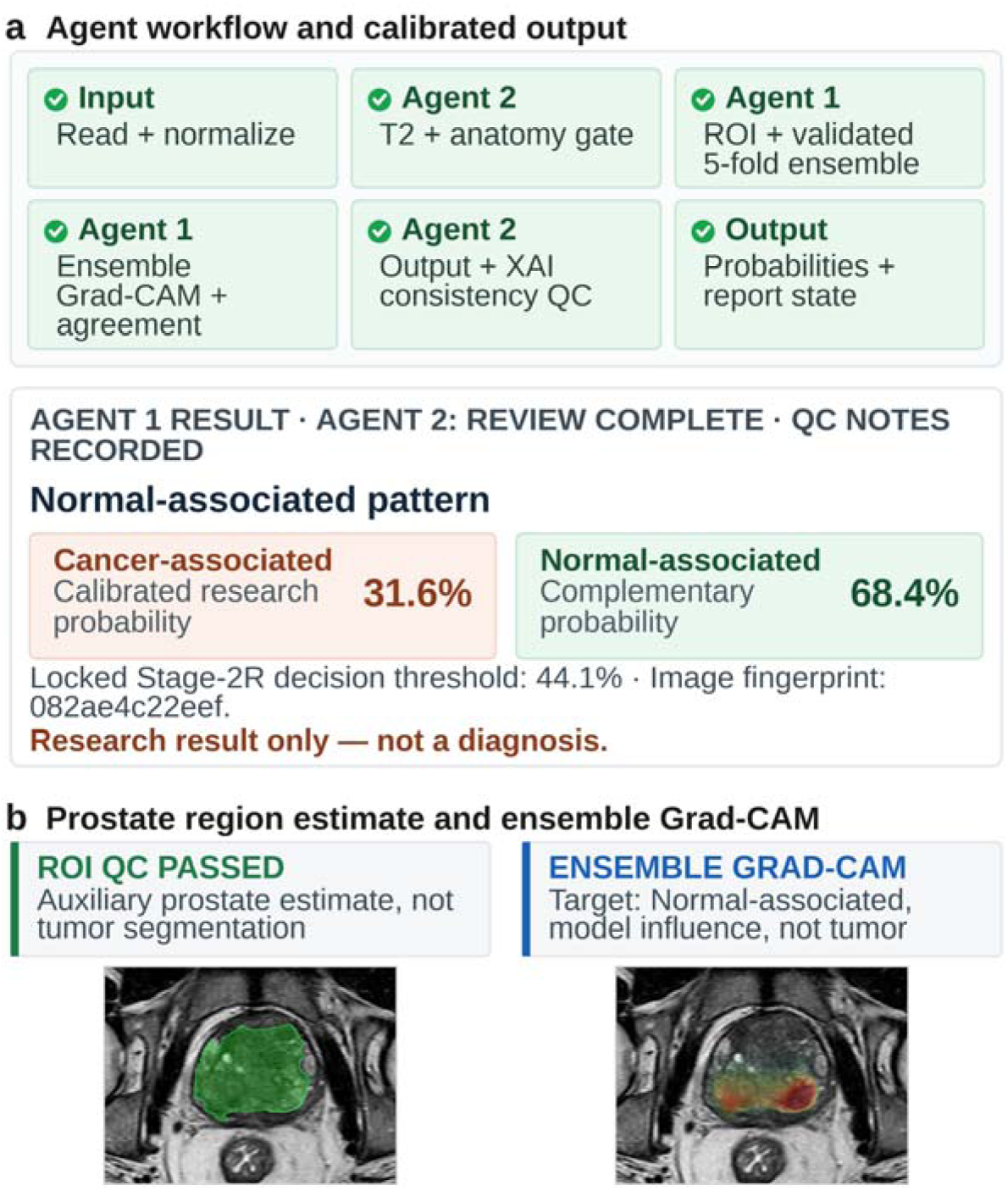
One case processed by the deployed dual-agent application. **(a)**, Fixed agent sequence and returned result; the quality agent recorded notes without changing the five-fold probability, calibration or threshold. **(b)**, Green marks the auxiliary whole-gland estimate, not a tumour segmentation; red to yellow marks stronger influence on the displayed class in the five-fold averaged Grad-CAM map, not a tumour boundary. Interface text is redrawn at legible size from the application output; probabilities, threshold, identifier and images are unchanged.

## Discussion

Four linked analyses answer the question posed at the outset. With 243 images and no diffusion data, out-of-fold discrimination reached 0.883. A separately specified volume model reached AUC 0.717 to 0.733 in a non-overlapping held-out cohort of 133 biopsy-linked patients, and an earlier frozen version 0.618 in 561 independent PROMIS patients. Distillation reproduced 77.6% of the frozen teacher’s probability variance, a transparent model 67.8%, and targeted interventions linked the output to gland size, low T2 signal and texture. Together these analyses show that T2-weighted MRI contains learnable cancer-related signal under limited-data conditions, that cross-domain performance remains moderate, and that the model’s computational dependencies can be audited.

Relative to our earlier slice-level framework, the present study advances the unit of analysis to the patient volume, shifts most capacity to frozen self-supervised^10^ and T2-specific features, and locks models, calibration and thresholds before evaluation. The final model achieved AUC 0.717 in the prespecified held-out analysis and 0.733 with exploratory test-time augmentation, and a separately frozen precursor retained above-chance discrimination in PROMIS at 0.618. These estimates describe the evolution and cross-domain behavior of the framework rather than one checkpoint across all cohorts.

Internal performance could be pushed higher by fitting and reporting on the same cohort, choosing a favorable operating point, or tuning against evaluation labels, none of which would be visible in the final figures. Internal predictions were generated out of fold; Prostate158 was reused during exploratory model development. The gap between the internal estimate and the held-out analysis, and the further fall in the independent cohort, is the cost of that discipline. A published T2-only model reported strong internal performance, but its healthy-control negatives, manual gland segmentation and random internal split differ materially from our clinically sourced labels and pooled out-of-fold evaluation^17^; no ranking is implied.

The PROMIS analysis provides the strongest evidence about independent transportability, although it evaluates a precursor, not the final checkpoint. Without retraining, fine-tuning, recalibration or threshold adjustment, the locked precursor retained above-chance discrimination for both any cancer and Gleason score 7 or higher under a different scanner fleet, protocol, reference standard and case mix. The AUC of 0.618 is therefore an independent framework-level stress test, not external validation of the final model, and its attenuation identifies protocol harmonization and prospective multicenter evaluation as the next requirements. The exploratory Prostate158 evaluation points the same way: above chance, far below the internal estimate.

The interpretability layer is computational rather than biological (Fig. 6). Attention weights were influential inside the frozen model, the strongest concepts tracked T2 signal and texture, and the boosting machine, model tree and rule tree exposed the same gland-size dependence; because the boosting machine is an exact additive decomposition, a single case can be explained by the signed terms that produced its score. The audit also revealed a weakness no accuracy metric would show, a rise in predicted risk when images are blurred, which shaped the quality gate. These computational explanations make model sensitivities inspectable and identify potential failure modes for subsequent clinical evaluation^8^.

The potential clinical role is specific and needs prospective evaluation. When diffusion images are degraded by metal or gas^4,5^, or legacy archives hold T2 only, a validated T2-only model could supply a research or quality-assurance signal where there is now none. Decision-curve analysis showed net benefit above biopsy-all and biopsy-none across a broad range. In the held-out cohort PPV was 0.946 but NPV only 0.271, largely reflecting 85% prevalence, so the model must not be used to exclude cancer. Realistic uses are retrospective archive analysis and image-quality auditing. In our earlier reader study, radiologists interpreting T2 alone achieved mean accuracy 82.7%, sensitivity 67.5% and Fleiss κ 0.524^12^ (Supplementary Table 4); because the reader and model evaluations were not paired, these results contextualize the difficulty of T2-only interpretation rather than establish model superiority.

Limitations define the next steps. The institutional unit of analysis is the image; patient-level linkage was unavailable. Consequently, patient overlap across internal folds cannot be excluded. The internal and held-out analyses use two models with different inputs, so this is a framework rather than one network. The PROMIS analysis evaluated a precursor under a different case mix, so it bounds cross-domain behavior rather than measuring the final model. The held-out interval is wide because only 20 patients had benign biopsies, and the interpretability cohort carries radiological rather than pathological labels. A prospective, paired reader study, with and without the model and with and without diffusion images, is the next step.

In summary, a widely acquired MRI sequence supported data-efficient prostate cancer classification, retained moderate discrimination in a held-out biopsy-linked cohort, showed attenuated but above-chance discrimination in an independent cohort, and could be audited concept by concept. Positioned as a transparent fallback and audit tool rather than a replacement for multiparametric MRI, it offers a practical route where resources are limited.

## Methods

### Ethics and data sources

This study was approved by the Ethics Committee of Golestan University of Medical Sciences (protocol IR.GOUMS.REC.1401.431) and performed in accordance with the Declaration of Helsinki and its later amendments. Informed consent was obtained from the participants. Public cohorts (PI-CAI^7^, Prostate158^14^, Prostate-MRI-US-Biopsy^13^, fastMRI Prostate^15^) were accessed through their official repositories under their licenses. Reporting follows TRIPOD+AI^18^.

### Institutional images and preprocessing

De-identified axial T2-weighted prostate MRI images collected at 5th Azar Hospital, Golestan University of Medical Sciences, Gorgan, Iran, between 2023 and 2026 were supplied in separate cancer and normal folders with the institutional label. Exact duplicates were identified by SHA-256 hash, with no conflicting-label duplicates. Images with more than 0.1% of pixels showing color differences above 12 grey levels between channels were treated as annotated and excluded, and the remainder converted to grayscale, resized with preserved aspect ratio and zero-padded to 224 × 224 pixels. A U-Net trained on PI-CAI whole-gland masks (validation Dice 0.935) supplied prostate masks; masks covering 1.5–60% of the image defined a bounding box with an 18% margin for region-of-interest input.

### Image-level classifier

Three backbones (ConvNeXt-Tiny, EfficientNet-B0, ResNet-50) were pre-trained for three epochs on 4,500 axial T2 slices from 1,500 PI-CAI examinations. Evaluation used five stratified outer folds. Within each training fold, 18% of images formed a validation split and a Tree-structured Parzen Estimator ran five trials over backbone, input mode, learning rate, weight decay and dropout, maximizing AUC + 0.20 × average precision − 0.10 × Brier score; the selection was trained for up to 12 epochs with AdamW and early stopping. Outer-fold predictions were pooled, Platt-scaled and thresholded at the Youden index (0.441), which was not transferred to the patient-level analyses. Confidence intervals used 2,000 bootstrap resamples. Net benefit was sensitivity × prevalence − (1 − specificity) × (1 − prevalence) × t/(1 − t).

### Volume model and patient-level evaluation

Each MRI volume was a bag of 12 axial T2 slices encoded by a frozen DINOv2 ViT-S/14 network^10^ into 384-dimensional vectors and pooled with gated attention^11^. Five fold models were trained on 400 UCLA (304 cancer, 96 benign) and 139 Prostate158 patients (83 with annotated tumor, 56 without), their mean calibrated by Platt scaling. A non-overlapping subset of 133 UCLA patients with at least six biopsy cores was fixed by SHA-256 manifest before fitting and excluded from training, calibration and threshold or model selection. Any Gleason pattern in any core defined cancer; the threshold was fixed at 0.50, confidence intervals came from bootstrap resampling, and 16-view test-time augmentation was applied to the same frozen model.

For independent external evaluation, a separately locked precursor version of the volume-model pipeline was applied once to 561 PROMIS patients^1^. All eligible patients were included, one prediction each, with checkpoint, calibration and threshold fixed beforehand and predictions fingerprinted before outcomes were read. Endpoints were any biopsy-confirmed cancer and Gleason score 7 or higher.

### Image-level model evaluated on Prostate158

A third model, used only for the exploratory Prostate158 evaluation, was trained on the 243 institutional images alone. Each image was described by intensity, grey-level co-occurrence, local binary pattern, gradient, Gabor, wavelet and gland-geometry descriptors, taken over the whole image and over a prostate region found by an unsupervised localizer. A soft-voting ensemble of a regularized logistic model, a random forest and a gradient-boosting model was fitted, regularization chosen inside training folds, and applied once to 137 Prostate158 patients. Because several configurations were compared on that cohort, the estimate is exploratory and optimistic.

### Computational interpretability and distillation

The explanation cohort was the fastMRI Prostate development split (264 patients), locked by hash. Each examination was described by attention summaries, intensity percentiles, grey-level co-occurrence texture, entropy, edge density, Laplacian variance, gland area and span, and activations of a 48-unit sparse autoencoder over the slice embeddings. Surrogates predicted the teacher probability or logit, never the outcome, under five-fold nested cross-validation, all selection confined to training folds. The eleven configurations ranged from a neural bag student and its concept blend to an explainable boosting machine, a model tree and a regression tree; fidelity was scored by *R*², mean absolute error, Pearson correlation and agreement at 0.50 (Supplementary Table 2). Interventions comprised zero-ablation of each concept, removal of the highest- and lowest-attention slice and directional concept sensitivity in 80 cases, and blur, contrast reduction and CLAHE in 40 cases. These describe the computations of a frozen model and are not evidence of biological causality.

### Application and reproducibility

An agent here is a bounded software component with explicit state, permitted actions, abstention behavior and auditable outputs; it does not update weights, change thresholds or learn online (Fig. 7). Manifests, checkpoints and prediction files were fingerprinted with SHA-256, and seeds were fixed; Prostate158 analyses involved exploratory configuration selection. Internal, held-out and independent external estimates are reported separately.

## Data availability

De-identified derived results, locked prediction files, evaluation code and model documentation are available at https://github.com/VahidMonfared/prostate-cancer-mri-ai. Institutional images are not publicly available because the analytic exports do not retain patient-level linkage or complete acquisition metadata and remain subject to institutional privacy and ethics restrictions. Public datasets are available from their providers: PI-CAI (https://pi-cai.grand-challenge.org), Prostate158 (https://github.com/kbressem/prostate158), Prostate-MRI-US-Biopsy (https://doi.org/10.7937/TCIA.2020.A61IOC1A) and fastMRI Prostate (https://fastmri.med.nyu.edu).

## Code availability

Code, model cards and trained weights are available at https://github.com/VahidMonfared/prostate-cancer-mri-ai. The dual-agent research application is available at https://huggingface.co/spaces/VahidMonfared/ProstateCancerAgent.

## Acknowledgements

We thank the teams that created and shared the PI-CAI, Prostate158, Prostate-MRI-US-Biopsy and fastMRI Prostate datasets. This work received no specific funding.

## Author contributions

V.M. conceived the study, developed and validated the models, performed the analyses, and prepared the initial manuscript draft. M.H.G. provided the clinical and radiological expertise, contributed the institutional imaging data, and advised on the medical and radiological aspects of the study. R.R. supervised the project, contributed methodological and conceptual guidance, and reviewed and edited the manuscript. All authors read and approved the final manuscript.

## Appendix

## Supplementary Information

This supplement contains three supplementary notes, four supplementary tables and seven supplementary figures, each with a short interpretation. Figure text is set at 16 pt, except the screenshots of the public application and the two curve panels exported by the locked analysis run, which are reproduced at their native resolution.

## Supplementary Note 1. Independent internal model

A hybrid model combined the image-level branch with a nine-slice, prostate-centered attention branch using nested cross-fitted weights (0.54 and 0.46). On the same 243 institutional images its nested out-of-fold performance was AUC 0.881, average precision 0.853, accuracy 0.802, sensitivity 0.890, specificity 0.720, PPV 0.750, NPV 0.874 and F1 0.814. Interpretation: a second, architecturally different model reproduces the internal result to within 0.002 AUC and trades specificity for sensitivity, which indicates that the internal estimate reflects the information in the images rather than one modelling choice. This model was retained for development only.

**Supplementary Table 1.** Configuration selected in each outer fold of the image-level classifier.

| Fold | Backbone | Input mode | Dropout | Epoch | Fold AUC |
| --- | --- | --- | --- | --- | --- |
| 0 | ConvNeXt-Tiny | Prostate region | 0.25 | 5 | 0.900 |
| 1 | ConvNeXt-Tiny | Prostate region | 0.10 | 11 | 0.960 |
| 2 | ResNet-50 | Whole image | 0.10 | 10 | 0.997 |
| 3 | ConvNeXt-Tiny | Prostate region | 0.40 | 12 | 0.883 |
| 4 | ConvNeXt-Tiny | Whole image | 0.40 | 10 | 0.917 |

Interpretation: the search converged on the prostate-region input and ConvNeXt-Tiny in four of five folds, and every fold exceeded an AUC of 0.88, so the internal result does not depend on one fortunate configuration. Fold sizes were 49, 49, 49, 48 and 48 images, and all backbones were initialized from PI-CAI T2 pre-training.

**Supplementary Fig. 1.**
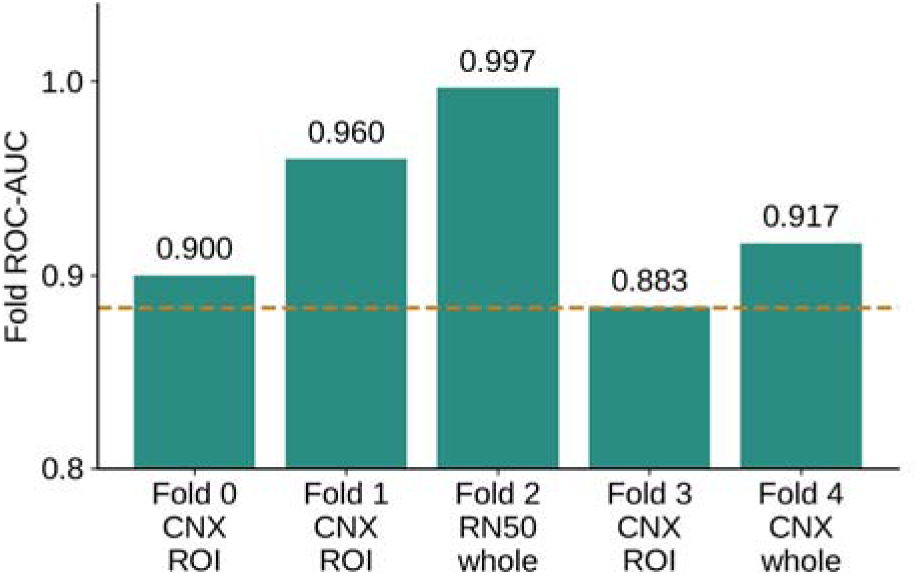
Fold-level internal ROC-AUC. Each bar is the AUC of one outer-fold model on its own held-out images, labelled with the selected backbone (CNX, ConvNeXt-Tiny; RN50, ResNet-50) and input mode. The dashed line marks the pooled out-of-fold AUC of 0.883. Interpretation: all five folds exceed 0.88, so internal performance is stable across data splits; the pooled estimate is lower than the fold mean because it combines probabilities from five different models, and it is the conservative figure we report.

**Supplementary Fig. 2.**
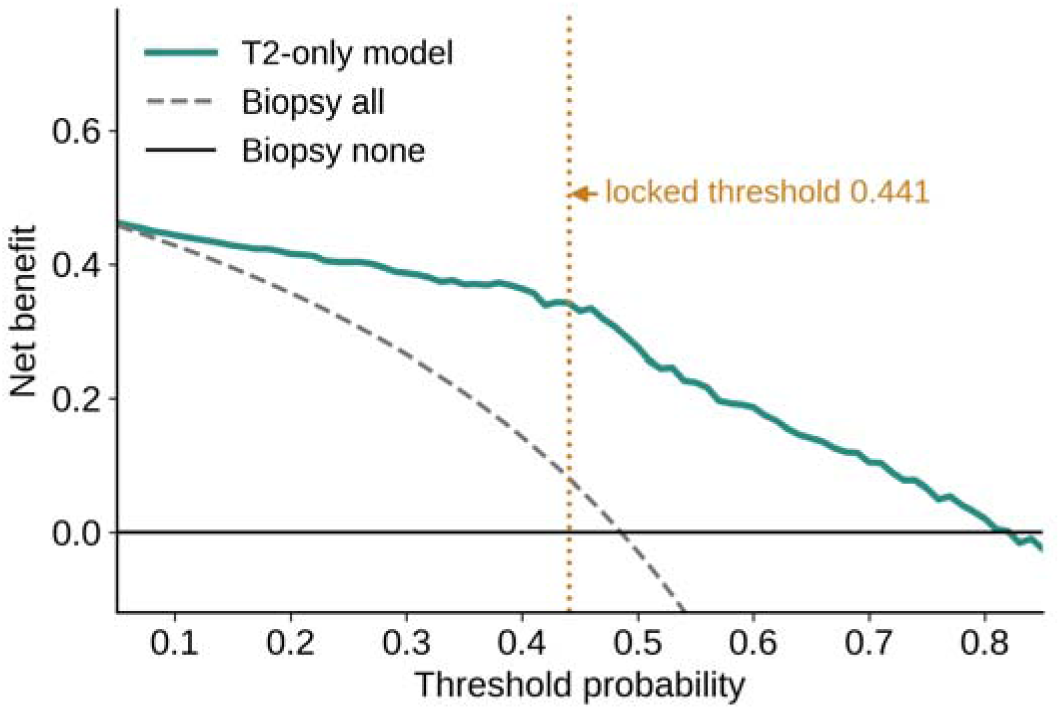
Decision-curve analysis of the internal out-of-fold predictions. Net benefit of using the model to select cases for further work-up, compared with treating all cases as positive (dashed) or none (solid black), across threshold probabilities. Interpretation: the model gives higher net benefit than both default strategies from about 0.05 to about 0.80, including the locked internal threshold of 0.441 (dotted line), which indicates usefulness across the plausible decision range rather than at a single cut-off.

**Supplementary Fig. 3.**
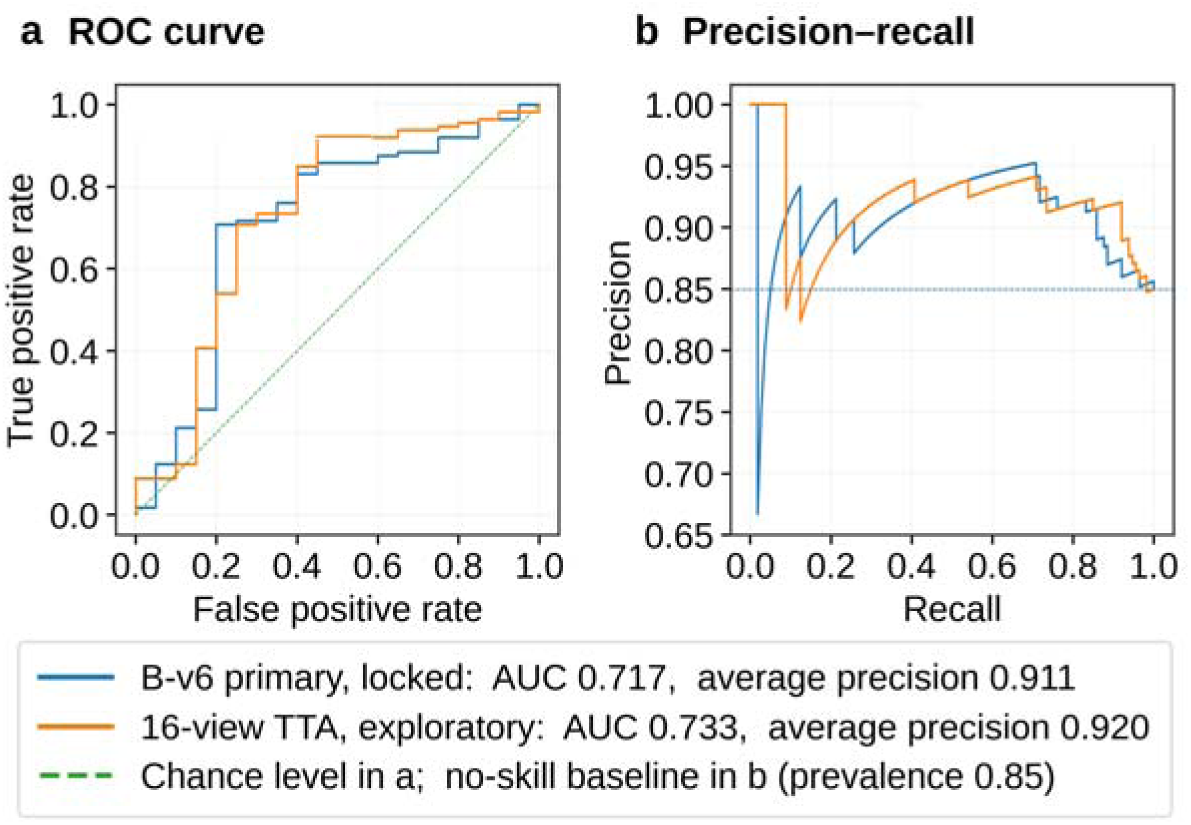
Discrimination curves for the locked volume model in the held-out cohort. **(a)**, Receiver operating characteristic and **(b)**, precision–recall curves for the 133 held-out patients, for the prespecified analysis and the 16-view test-time augmentation of the same frozen model. Interpretation: the curves rise steeply at high specificity, where the fixed 0.50 operating point sits, and precision stays above 0.90 for most of the recall range, consistent with the reported positive predictive value of 0.946. Both panels are reproduced from the locked analysis run.

**Supplementary Fig. 4.**
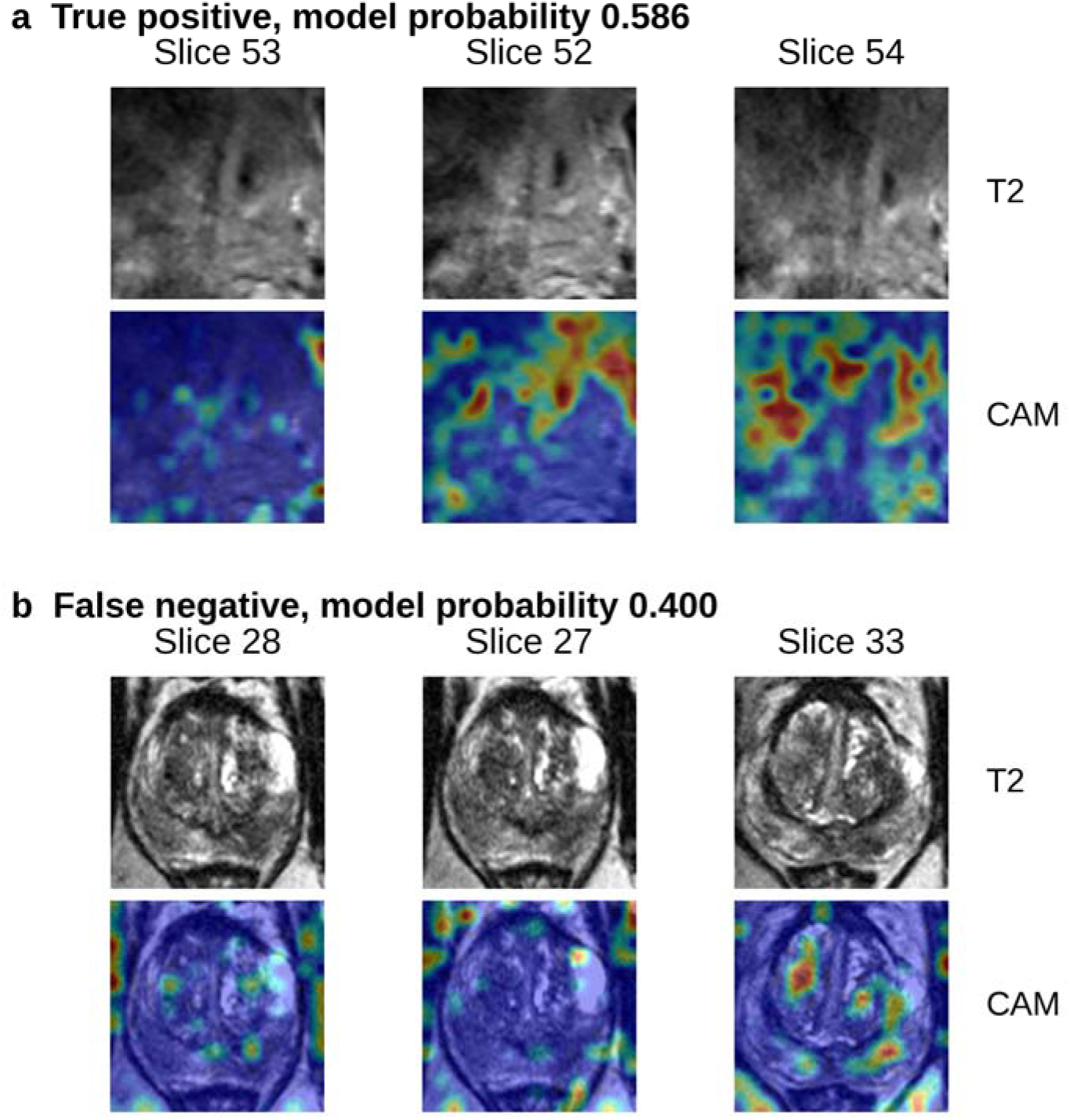
Attribution maps for a correct and an incorrect held-out case. Transformer-adapted Grad-CAM for the three highest-attention slices in a, a true positive and b, a false negative from the held-out cohort, with the T2 slice above and the map below. Interpretation: in the true positive the strongest response lies in the posterior peripheral zone, whereas in the missed cancer the response is diffuse with no dominant focus, the visual signature of an uncertain case. Maps show where the frozen model was influenced; they are attribution and quality-control outputs, not tumor segmentations.

**Supplementary Table 2.**
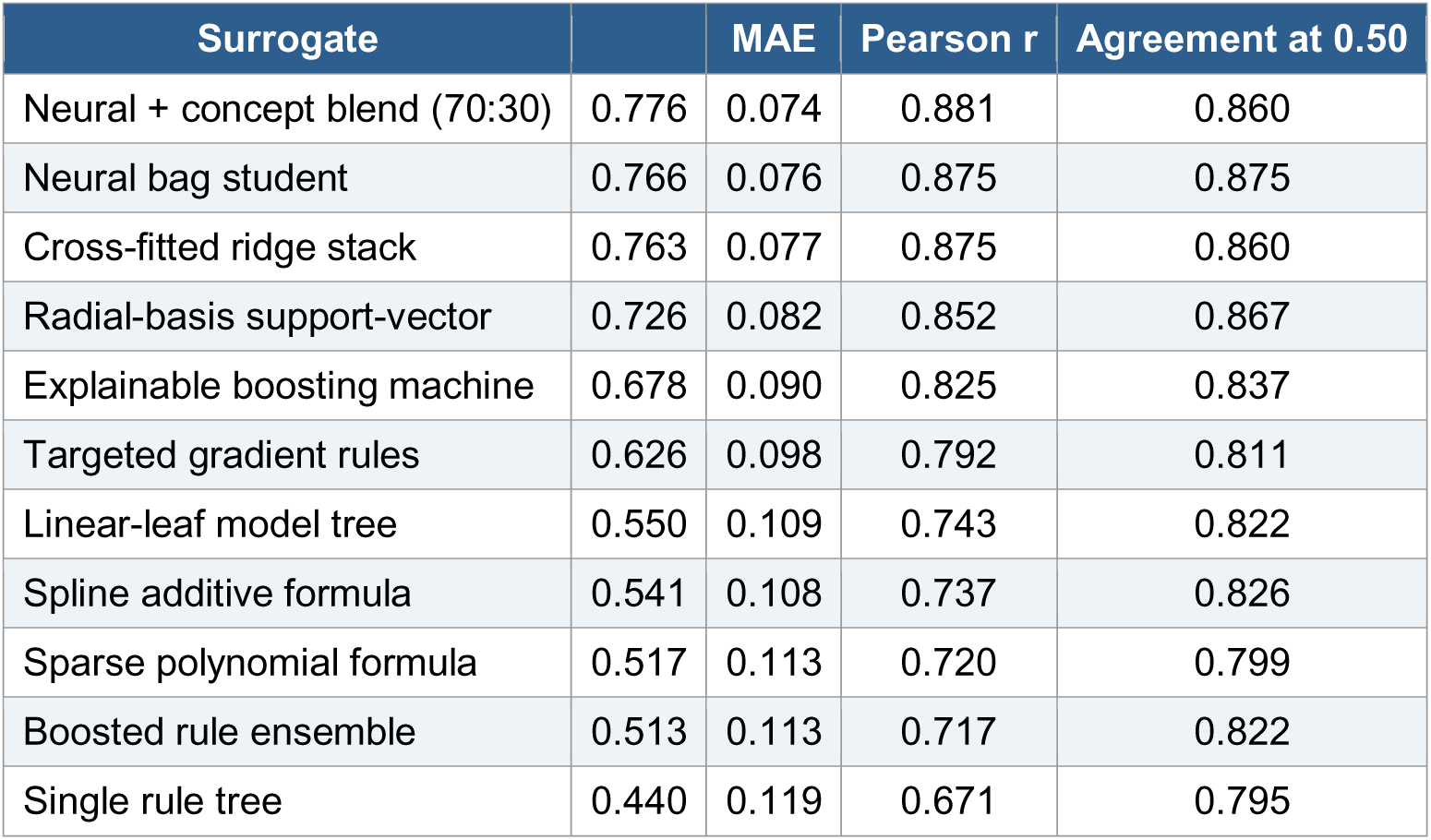
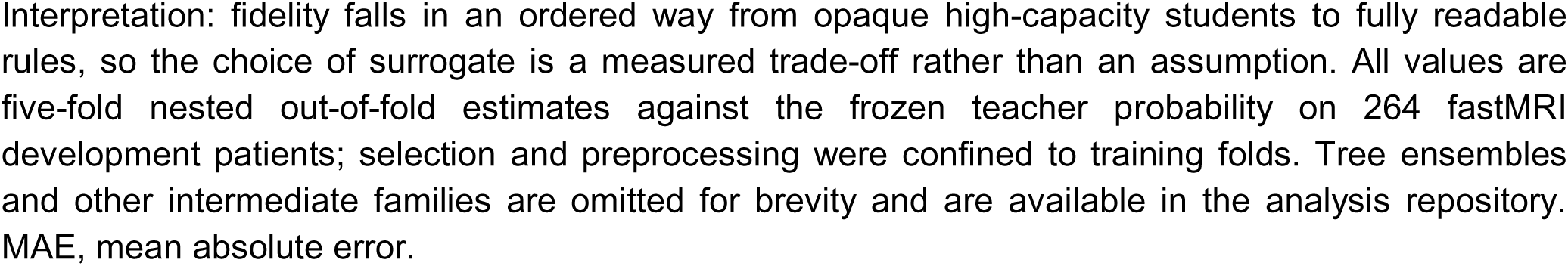
Fidelity of every surrogate family to the frozen teacher.

| Surrogate |  | MAE | Pearson r | Agreement at 0.50 |
| --- | --- | --- | --- | --- |
| Neural + concept blend (70:30) | 0.776 | 0.074 | 0.881 | 0.860 |
| Neural bag student | 0.766 | 0.076 | 0.875 | 0.875 |
| Cross-fitted ridge stack | 0.763 | 0.077 | 0.875 | 0.860 |
| Radial-basis support-vector | 0.726 | 0.082 | 0.852 | 0.867 |
| Explainable boosting machine | 0.678 | 0.090 | 0.825 | 0.837 |
| Targeted gradient rules | 0.626 | 0.098 | 0.792 | 0.811 |
| Linear-leaf model tree | 0.550 | 0.109 | 0.743 | 0.822 |
| Spline additive formula | 0.541 | 0.108 | 0.737 | 0.826 |
| Sparse polynomial formula | 0.517 | 0.113 | 0.720 | 0.799 |
| Boosted rule ensemble | 0.513 | 0.113 | 0.717 | 0.822 |
| Single rule tree | 0.440 | 0.119 | 0.671 | 0.795 |

Interpretation: fidelity falls in an ordered way from opaque high-capacity students to fully readable rules, so the choice of surrogate is a measured trade-off rather than an assumption. All values are five-fold nested out-of-fold estimates against the frozen teacher probability on 264 fastMRI development patients; selection and preprocessing were confined to training folds. Tree ensembles and other intermediate families are omitted for brevity and are available in the analysis repository. MAE, mean absolute error.

**Supplementary Fig. 5.**
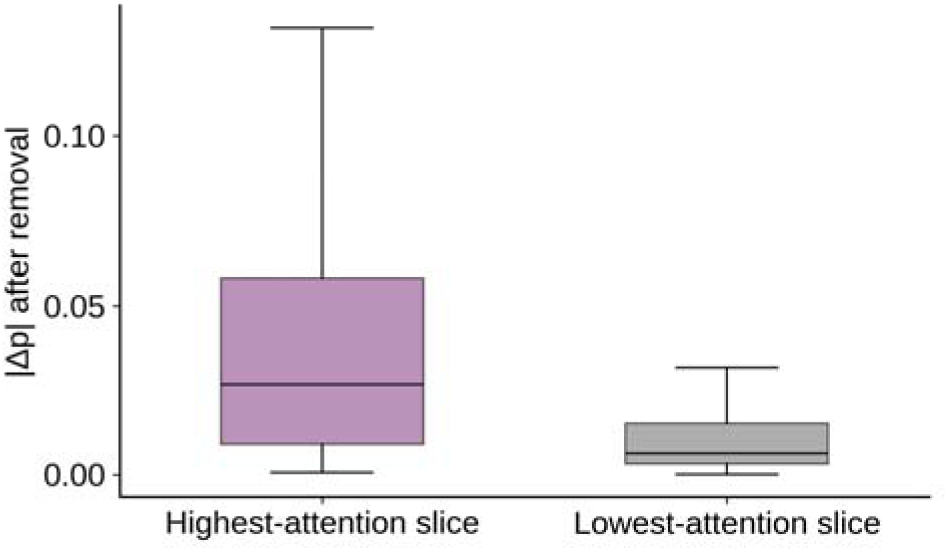
Attention-slice ablation in the frozen teacher. Absolute change in teacher probability after removing the highest-attention or the lowest-attention slice from each bag (80 cases; boxes show median, interquartile range and 1.5 × IQR whiskers). Interpretation: removing the slice the model attends to changes the output more than three times as much as removing the least-attended slice (means 0.040 versus 0.012; paired Wilcoxon ), so the displayed attention is a faithful pointer to the slices that drive the score.

**Supplementary Table 3.** Most influential sparse-autoencoder concepts.

| Concept | Mean $\vert \Delta p\vert $ | Max $\vert \Delta p\vert $ | Strongest descriptor (r) | Second descriptor (r) |
| --- | --- | --- | --- | --- |
| C38 | 0.148 | 0.363 | 10th-percentile intensity (−0.37) | 10th-percentile maximum (−0.30) |
| C28 | 0.108 | 0.293 | GLCM contrast (−0.60) | GLCM homogeneity, max (0.50) |
| C32 | 0.102 | 0.251 | Selected slice position (−0.37) | Gland area (−0.35) |
| C23 | 0.102 | 0.246 | GLCM contrast (−0.35) | Mean intensity (−0.32) |
| C04 | 0.100 | 0.332 | GLCM contrast (−0.42) | GLCM homogeneity SD (0.40) |
| C30 | 0.099 | 0.350 | 10th-percentile intensity (0.22) | Laplacian variance (0.19) |
| C42 | 0.097 | 0.367 | 10th-percentile intensity (0.40) | 10th-percentile maximum (0.34) |
| C11 | 0.095 | 0.271 | Edge density SD (0.51) | GLCM energy, max (0.50) |

**Interpretation:** the concepts that move the model most are intensity and texture descriptors, headed by low-percentile signal and grey-level co-occurrence (GLCM) contrast, which is what radiologists use when they call a T2 lesion hypointense and homogeneous. Zero-ablation was performed in 80 fastMRI development cases; descriptor correlations state what each concept tracks in the images and do not establish biological meaning.

**Supplementary Fig. 6.**
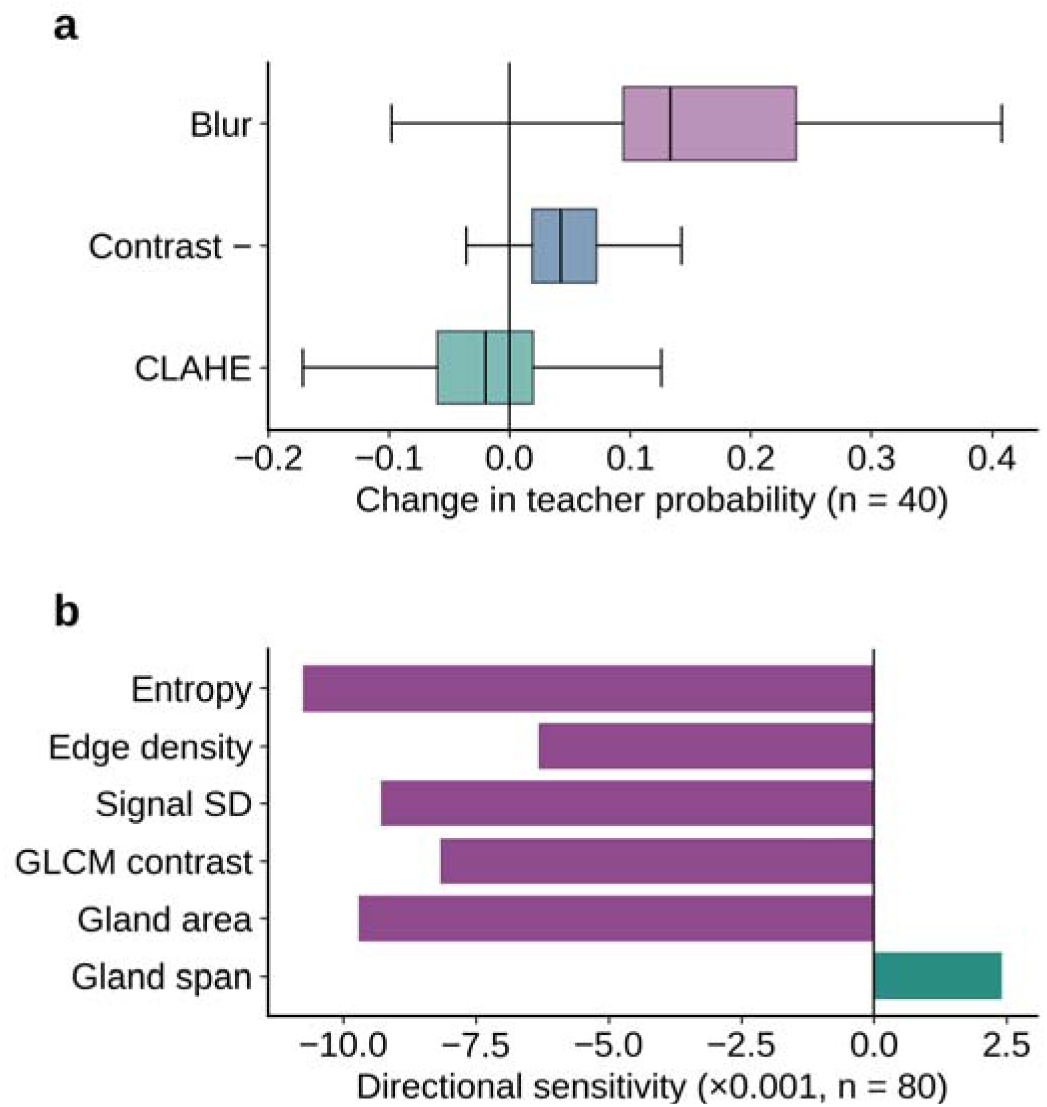
Input interventions and concept sensitivity. **(a)**, Change in teacher probability after blurring, contrast reduction and contrast-limited adaptive histogram equalization (CLAHE) in 40 development cases (mean +0.167, +0.051 and −0.022); boxes show median and interquartile range. **(b)**, Mean directional sensitivity to six engineered concepts in 80 cases. Interpretation: removing fine texture by blurring raises predicted risk, so image sharpness must be checked before a score is trusted, which is what the quality agent does; sensitivity was negative in all 80 cases for entropy, edge density, signal standard deviation, grey-level co-occurrence contrast and gland area, and positive in 79 of 80 for gland span, indicating a consistent direction of effect.

## Supplementary Note 2. Reading the interpretable models

The reported surrogate is the fixed 70:30 blend of the neural bag and concept students (Equation 1). The explainable boosting machine (Equation 2) stores one learned shape function per feature and twenty pairwise interaction surfaces, so the score of an individual patient is the intercept plus a short list of signed contributions that can be printed beside the image; the fitted object and its effect curves are released with the analysis code. The compact model tree (Equation 3) applies one of two ridge equations according to mean segmented gland area, and the single auditable rule tree expands the same structure into ten explicit rules (Fig. 5b). All three describe the computations of the frozen teacher and are not claims about biology; none of them replaces the classifier, whose probabilities remain the reported output.

**Supplementary Fig. 7.**
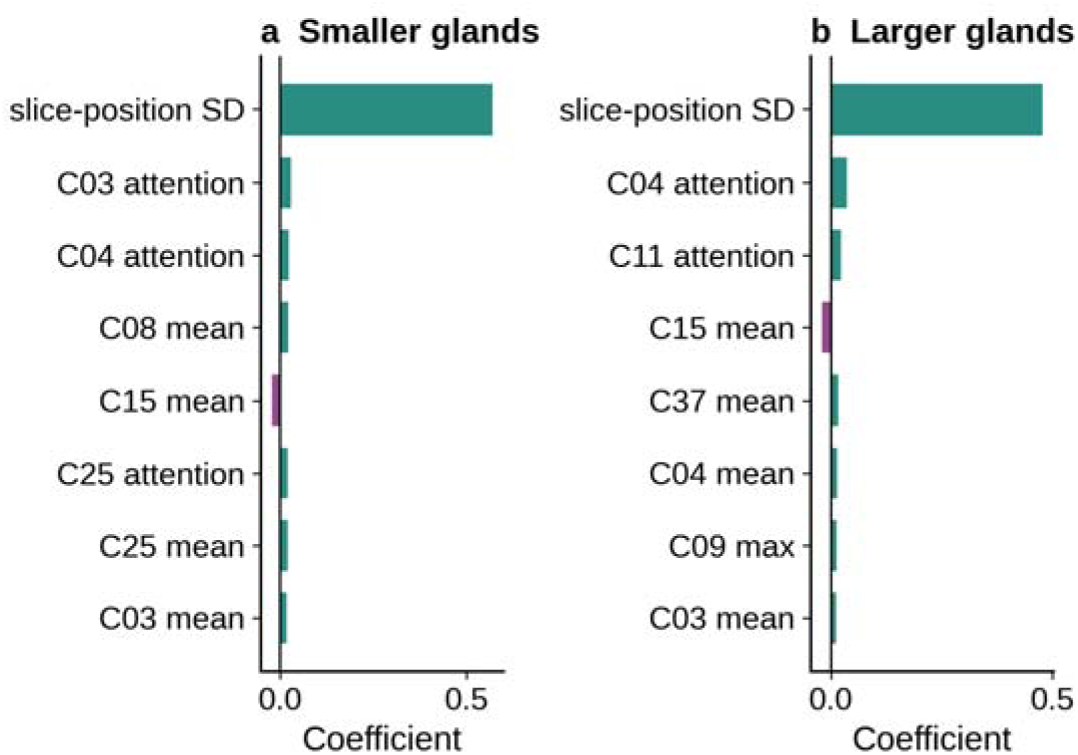
Leaf equations of the compact model tree. The eight largest coefficients of the ridge equation fitted inside each branch of the one-split model tree (Equation 3): **(a)**, smaller glands ( , 91 patients); **(b)**, larger glands ( , 173 patients). Interpretation: both branches are dominated by the spread of the selected slice positions, an acquisition-related quantity, followed by sparse-autoencoder concept activations, and the two branches weight the same concepts differently, which is why one linear equation fitted to all patients is less faithful than the split model. The complete 24-term equations are provided with the analysis code.

Equation 1 combines two student predictions; Equation 2 decomposes the EBM prediction into feature and interaction contributions; Equation 3 selects a branch-specific linear predictor according to gland area.

## Supplementary Note 3. Exploratory external evaluation on Prostate158

Because the volume model was developed partly on Prostate158 patients, that collection cannot serve as an external test for it. We therefore trained a separate image-level model on the 243 institutional images alone, using intensity, grey-level co-occurrence, local binary pattern, wavelet and gland-geometry descriptors computed after an unsupervised gland localization step, and applied it once to 137 Prostate158 patients (82 with tumor annotation, 55 without), one canonical axial T2 image each. Patient-level ROC-AUC was 0.643 (95% CI 0.545–0.734), average precision 0.728, with sensitivity 0.951, specificity 0.146, accuracy 0.628 and F1 0.754 at the internally locked threshold; at a post-hoc operating point specificity rose to 0.582 with sensitivity 0.695 and positive predictive value 0.713. Its five-repeat nested internal estimate on the institutional images was AUC 0.924 (0.889-0.953). Interpretation: this is the first evaluation in this line of work whose confidence interval excludes chance, against slice-level AUCs of 0.416 to 0.494 for six configurations reported previously. It is exploratory rather than confirmatory, because several feature representations, domain-alignment methods and aggregation rules were compared on this cohort before the reported value was chosen, so the estimate is optimistic, and no formal comparison should be drawn from it.

**Supplementary Table 4.** Prior T2-only reader study.

| Readers and cases | Sensitivity | Specificity | Accuracy | Agreement |
| --- | --- | --- | --- | --- |
| Five board-certified radiologists; 22 cases (8 cancer, 14 normal) | 67.5% (mean) | 91.4% (mean) | 82.7% (mean) | Fleiss $\kappa$ 0.524 (0.314–0.734) |

Interpretation: experts reading T2 alone were specific but missed about a third of cancers and agreed only moderately, which is the clinical gap this work addresses. Data from ref. <sup>12</sup>; the reader cases do not overlap the cohorts analyzed here, so no paired comparison is made.

## References

1. Ahmed, H. U. et al. Diagnostic accuracy of multi-parametric MRI and TRUS biopsy in prostate cancer (PROMIS): a paired validating confirmatory study. Lancet 389, 815–822 (2017).

2. Kasivisvanathan, V. et al. MRI-targeted or standard biopsy for prostate-cancer diagnosis. N. Engl. J. Med. 378, 1767–1777 (2018).

3. Turkbey, B. et al. Prostate Imaging Reporting and Data System version 2.1: 2019 update of Prostate Imaging Reporting and Data System version 2. Eur. Urol. 76, 340–351 (2019).

4. Nakai, H. et al. Decreased prostate MRI cancer detection rate due to moderate to severe susceptibility artifacts from hip prosthesis. Eur. Radiol. 34, 3387–3399 (2024).

5. Nakai, H. et al., Prostate MRI cancer detection rate by deep learning-assisted image quality categorization: gas-induced susceptibility artifacts in diffusion-weighted imaging. Insights Imaging 16, 217 (2025).

6. Hosseinzadeh, M. et al. Deep learning-assisted prostate cancer detection on bi-parametric MRI: minimum training data size requirements and effect of prior knowledge. Eur. Radiol. 32, 2224–2234 (2022).

7. Saha, A. et al. Artificial intelligence and radiologists in prostate cancer detection on MRI (PI-CAI): an international, paired, non-inferiority, confirmatory study. Lancet Oncol. 25, 879–887 (2024).

8. Rudin, C. Stop explaining black box machine learning models for high stakes decisions and use interpretable models instead. Nat. Mach. Intell. 1, 206–215 (2019).

9. Buciluǎ, C., Caruana, R. & Niculescu-Mizil, A. Model compression. In Proc. 12th ACM SIGKDD International Conference on Knowledge Discovery and Data Mining 535–541 (ACM, 2006).

10. Oquab, M. et al. DINOv2: learning robust visual features without supervision. Trans. Mach. Learn. Res. (2024).

11. Ilse, M., Tomczak, J. & Welling, M. Attention-based deep multiple instance learning. In Proc. 35th International Conference on Machine Learning, PMLR 80, 2127–2136 (2018).

12. Monfared, V., et al. Interpretable prostate cancer detection using a small cohort of MRI images. Preprint at https://arxiv.org/abs/2603.18460 (2026).

13. Natarajan, S., Priester, A., Margolis, D., Huang, J. & Marks, L. Prostate MRI and ultrasound with pathology and coordinates of tracked biopsy (Prostate-MRI-US-Biopsy), version 2. The Cancer Imaging Archive 10.7937/TCIA.2020.A61IOC1A (2020).

14. Adams, L. C. et al. Prostate158: an expert-annotated 3T MRI dataset and algorithm for prostate cancer detection. Comput. Biol. Med. 148, 105817 (2022).

15. Tibrewala, R. et al. FastMRI Prostate: a public, biparametric MRI dataset to advance machine learning for prostate cancer imaging. Sci. Data 11, 404 (2024).

16. Selvaraju, R. R. et al. Grad-CAM: visual explanations from deep networks via gradient-based localization. In Proc. IEEE International Conference on Computer Vision 618–626 (2017).

17. Jin, L. et al. T2-weighted imaging-based deep-learning method for noninvasive prostate cancer detection and Gleason grade prediction: a multicenter study. Insights Imaging 15, 111 (2024). 10.1186/s13244-024-01682-z

18. Collins, G. S. et al. TRIPOD+AI statement: updated guidance for reporting clinical prediction models that use regression or machine learning methods. BMJ 385, e078378 (2024).

